# One Size Fits All? Comparing Foundation and Task-specific Models for Retinal Fluid Segmentation

**DOI:** 10.64898/2026.04.26.26351792

**Authors:** Xiaoyu Sun, Saiyu You, Siqi Sun, Cindy X. Cai, Joanna Abraham, Po-Yin Yen, Linying Zhang

## Abstract

Retinal fluids, detectable through optical coherence tomography (OCT), are key biomarkers for retinal diseases such as diabetic macular edema and age-related macular degeneration, guiding treatment decisions and monitoring response to therapy. Automated segmentation of retinal fluids could support large-scale clinical research and the development of clinical decision support tools. Recent ophthalmic foundation models trained on massive retinal imaging datasets show promise across many downstream tasks, including disease risk prediction and biomarker segmentation, but their performance relative to task-specific models for specialized clinical tasks remains unclear. We compared a task-specific segmentation model (RetiFluidNet) and an ophthalmic foundation model (VisionFM) using a standard benchmarking dataset containing 4,248 OCT images from 48 patients with three retinal diseases. Models were evaluated using three-fold cross-validation and assessed for pixel-level segmentation accuracy and patient-level fluid burden estimation. The task-specific model achieved higher segmentation performance and more consistent fluid quantification across devices. These findings suggest that, for retinal fluid segmentation, specialized task-specific models currently remain more reliable than general-purpose foundation models, highlighting the need for targeted adaptation before clinical deployment.

## 1. Introduction

Foundation models pretrained on large-scale imaging data have emerged as a transformative paradigm in medical imaging, enabling models to learn generalizable visual representations that can be adapted to diverse downstream tasks.^1, 2^ Recent studies have shown that such models can improve performance in settings with limited labeled data and facilitate transfer across imaging modalities, diseases, and institutions.^2^ In ophthalmology, vision foundation models trained on millions of retinal images have demonstrated promising results across tasks such as disease classification, segmentation, and multimodal prediction.^5^ Their ability to leverage large unlabeled datasets and transfer learned representations to new tasks make them particularly attractive in medical imaging domains where expert annotations are costly and time-consuming to obtain.^2^ However, despite their broad capabilities, it remains unclear whether foundation models consistently match or surpass locally trained, task-specific models in highly specialized clinical applications that require fine-grained structural delineation.

Optical coherence tomography (OCT) is routinely used in clinical practice to diagnose and monitor retinal diseases, and quantitative assessment of retinal fluid plays a central role in guiding treatment decisions. Retinal fluid segmentation from OCT represents a particularly challenging and clinically important task because fluid pockets are often small, low-contrast, and embedded within complex retinal anatomy. We focus on three retinal biomarkers in macular OCT: intraretinal fluid (IRF), subretinal fluid (SRF), and pigment epithelial detachment (PED). These retinal biomarkers are widely used in the evaluation of retinal diseases such as diabetic macular edema (DME) and age-related macular degeneration (AMD) and are clinically relevant for treatment planning and monitoring response to anti-VEGF therapy.^3^ Reliable retinal biomarker segmentation from OCTs is therefore important not only for anatomical delineation of disease lesions but also for downstream quantification of retinal fluid burden, which can support longitudinal monitoring and treatment evaluation.

In this study, we performed a direct comparison between a task-specific model, RetiFluidNet (RFN), and a recently released foundation model in ophthalmology, VisionFM (VFM), for multi-class retinal fluid segmentation.^4, 5^ Using the MICCAI 2017 RETOUCH dataset of expert-annotated OCT volumes, where each volume represents a three-dimensional retinal scan composed of a series of two-dimensional cross-sectional images (B-scans) from one patient, we evaluate (1) pixel-level segmentation accuracy for IRF, SRF, and PED and (2) patient-level fluid burden estimates derived from segmentation outputs.^6^ This head-to-head evaluation is intended to clarify the relative strengths and limitations of task-specific versus foundation-model approaches for OCT retinal biomarker segmentation.

## 2. Materials and Methods

### 2.1. Data source

We used the MICCAI 2017 RETOUCH challenge dataset that includes OCTs acquired from two devices: Zeiss Cirrus and Heidelberg Spectralis (24 volumes from each device).^6^. The two OCT devices have different acquisition characteristics: each Cirrus volume contains 128 B-scans at 512×1024 pixels, yielding 3,072 B-scans for the Cirrus subset, while each Spectralis volume contains 49 B-scans at 512×496 pixels, yielding 1,176 B-scans for the Spectralis subset. In total, the dataset contains 4,248 B-scans. Pixel-level ground-truth annotations for intraretinal fluid (IRF), subretinal fluid (SRF), and pigment epithelial detachment (PED) were provided by multiple expert graders in the MICCAI challenge with consensus adjudication and served as the reference standard for all evaluations.^6^

### 2.2. Data preprocessing

All RETOUCH OCT volumes and their corresponding annotation masks were converted to per–B-scan PNG images to standardize file formats across devices while preserving alignment between each B-scan and its corresponding mask. To harmonize spatial resolution and field of view between devices and to meet a unified model input size, each B-scan was resized to 256×256 pixels. We used bilinear interpolation for OCT intensity images and nearest-neighbor interpolation for segmentation masks to avoid creating invalid label values. Masks represented four classes (background, IRF, SRF, PED). In the stored mask images, these classes were encoded as grayscale values {0, 85, 170, 255}, which we mapped to integer labels {0, 1, 2, 3} prior to training. Finally, each B-scan was independently min-max normalized to the range [0, 1] to reduce inter-scan intensity variability. This led to normalized 256×256 PNG images for B-scans and masks.

We augmented the training sets using standard data augmentation techniques to increase sample size, which has been shown to improve segmentation model.^7^ Data augmentation was applied only to training sets and included horizontal flips, contrast adjustments, small rotations (±0.01, ±0.02, ±0.05 radians), and translations, expanding the effective training dataset to 39,648 images. Masks were transformed using nearest-neighbor interpolation to preserve discrete labels. All input images were resized to 256×256 pixels. The overall study workflow is summarized in Figure 1.

**Figure 1.**
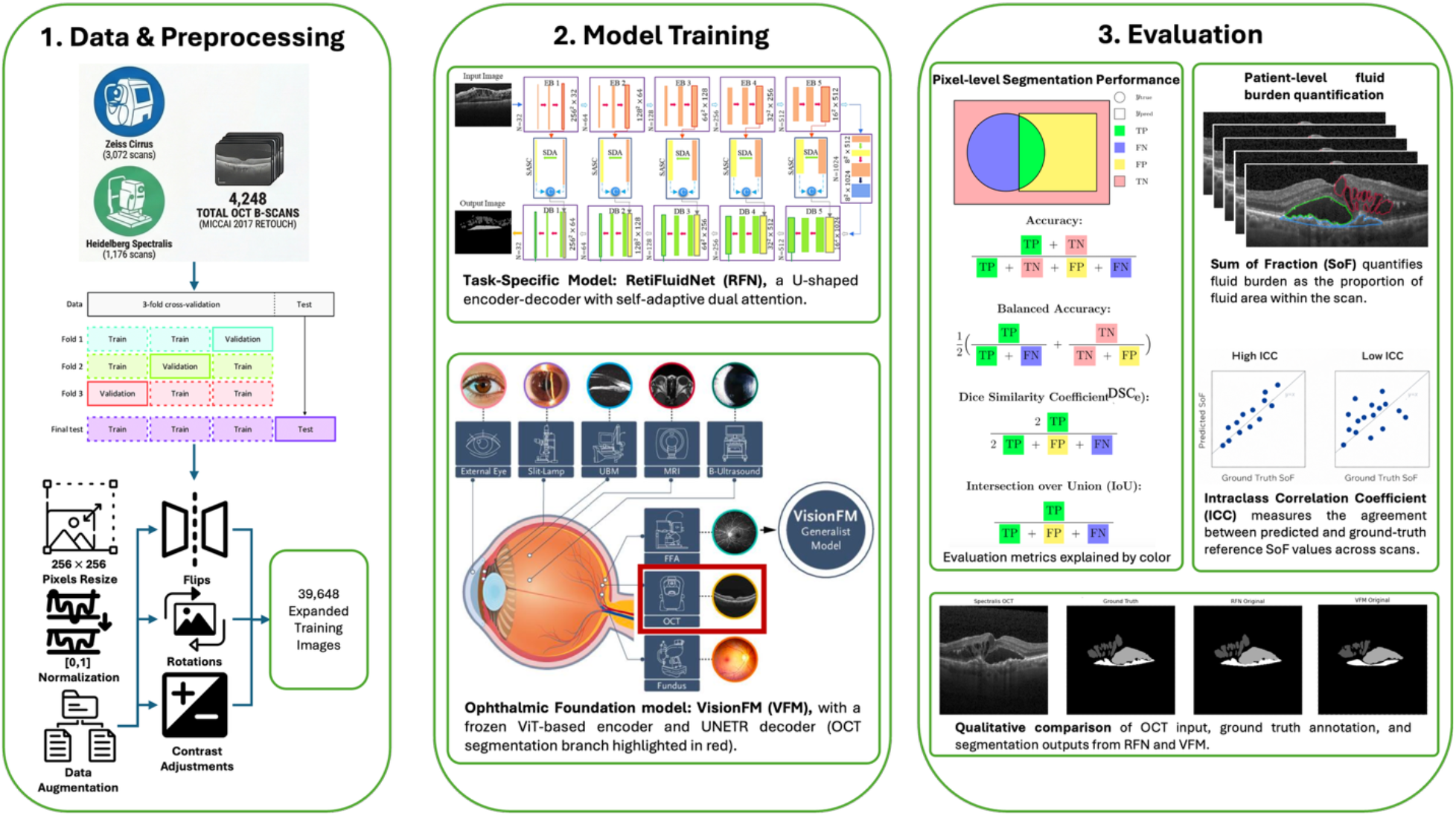
Schematic view of the study workflow. OCT volumes from the MICCAI 2017 RETOUCH dataset were preprocessed and split using three-fold, volume-level cross-validation. Two models, the task-specific RetiFluidNet (RFN) and the ophthalmic foundation model VisionFM (VFM), were trained and evaluated under identical preprocessing and cross-validation settings. Performance was assessed using pixel-level segmentation metrics, patient-level fluid burden estimates, and qualitative visual comparison of randomly sampled B-scans.

### 2.3. Model architecture and training workflow

The goal of this study was to evaluate the performance of task-specific models versus vision foundation models on segmentation performance. The project workflow was shown in Figure 1. The architecture and training of the two models were as follows:

#### Task-specific model

For the task-specific model, we adopted the architecture of a previously published model called RetiFluidNet (RFN).^4^ RFN was a deep learning model developed specifically for retinal fluid segmentation from OCTs. It adopts a U-shaped encoder–decoder architecture with convolutional layers as the backbone of the encoder and the decoder. RFN also employs a dual-attention module that captures the relevant contextual information in the spatial and channel domains. In this experiment, we trained RFN, including both the encoder and the decoder. The training objective is a joint loss that linearly combines multi-scale Dice-overlap terms (dice loss components; DLC) with an edge-preserving connectivity-based loss (connectivity loss components; CLC), where local connectivity masks are used to better model inter-pixel relationships and enhance boundary delineation for each fluid class.^4^

#### Foundation model

For ophthalmic foundation model, we included VisionFM (VFM).^5^ VFM is a multimodal ophthalmic foundation model pretrained on 3.4 million retinal images across 8 imaging modalities, including OCT modality, across 560,457 individuals. Architecturally, VFM adopts an encoder-decoder design: a ViT-Base transformer encoder pretrained with iBOT extracts hierarchical feature representations, which are then upsampled and fused by an UNETR-style segmentation decoder to produce pixel-wise masks.^8, 9, 10^ In this experiment, we applied the pretrained transformer-based encoder to extract embeddings for our OCTs, and then we trained only the segmentation decoder on RETOUCH labels, following VisionFM repository workflow for downstream decoder training. The decoder was optimized using a Dice–focal composite objective (DiceFocalLoss) to address class imbalance.^5, 11^

To ensure fair comparison, we conducted three-fold cross-validation using predefined volume-level splits, preventing data leakage across B-scans from the same OCT volume. Folds were stratified by device type to balance Cirrus and Spectralis scans. Both models were trained using hyperparameters reported in their original implementations. RFN was trained for up to 30 epochs using RMSprop with a per-GPU batch size of 4, an initial learning rate of 2×10^−4^, and step decay by a factor of 0.8 every 5 epochs. For VFM, we froze the pretrained OCT encoder and optimized only the UNETR-style segmentation decoder. VFM was trained for up to 30 epochs using AdamW with a per-GPU batch size of 64, an initial learning rate of 1×10^−3^, and weight decay of 0.05.

### 2.4. Evaluation criteria

We evaluated model performance both quantitatively and qualitatively at three levels: (1) pixel-level segmentation performance, (2) patient-level fluid burden quantification performance, and (3) image-level qualitative assessment through visualization.

#### Pixel-level segmentation performance

We report accuracy (ACC), balanced accuracy (BAcc), Dice similarity coefficient (DSC), and intersection over union (IoU) computed between the predicted segmentation mask *y*_*pred*_ and the ground-truth mask *y*_*true*_. Since our task is multi-class segmentation with four labels (background, IRF, SRF, and PED), we compute these metrics per class in a one-vs-rest manner. For a given class *c* (background, IRF, SRF, and PED), pixels labeled *c* are treated as positives and all other pixels as negatives, which defines true positives (*TP*), true negatives (*TN*), false positives (*FP*), and false negatives (*FN*) for that class. Background was included as a formal segmentation class to represent none-fluid regions, although the primary classes of clinical interest were IRF, SRF, and PED. Metrics were computed per B-scan and then averaged across B-scans within each fold.

ACC measures the proportion of correctly classified pixels but can be dominated by the background class in highly imbalanced segmentation, where predicting background everywhere can yield deceptively high ACC:

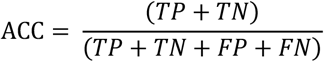

BAcc addresses this by averaging sensitivity (true positive rate) and specificity (true negative rate), reducing the influence of class prevalence and better reflecting performance on the minority class, which in our setting corresponds to fluid pixels:

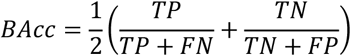

DSC quantifies spatial overlap between the predicted and ground-truth masks and is equivalent to the F1-score for the positive class. In segmentation settings with small structures, DSC is sensitive to false positives and false negatives introduced by small boundary misalignments:

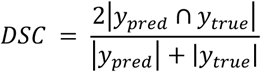

IoU computes the intersection over the union of predicted and ground-truth regions and is a more stringent overlap metric that penalizes extra predicted area and missed area more strongly:

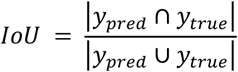

#### Patient-level fluid burden quantification performance

In addition to pixel-wise segmentation metrics, we quantified patient-level fluid burden using a pixel-based measure derived from the segmented OCT volume. For each patient *p* and fluid class *c* ∈{IRF, SRF, PED}, the total number of pixels labeled as class *c* across all *M*_*p*_ B-scans from a patient *p* can be denoted as

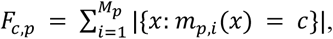

where *m*_*p,i*_(*x*) denotes the label at pixel *x* in the *i*-th B-scan for patient *p*. When voxel spacing metadata (i.e., the spacing between B-scans) are available, the 3D physical fluid volume can be calculated and is proportional to this total fluid pixel count *F*_*c,p*_, with the proportionality factor determined by voxel dimensions.^19, 20^ However, the RETOUCH dataset does not provide voxel spacing metadata, so the physical fluid volume cannot be directly computed. Therefore, we used the segmented fluid extent itself as a proxy measure of the 3D fluid volume.

To reduce sensitivity to image resolution, we normalized the fluid pixel count in each B-scan by the total number of pixels in a B-scan. We defined the sum of fractions (SoF) as

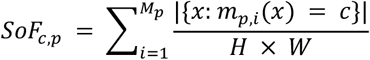

where *H* and *W* denote the height and width in pixels of a B-scan. Because OCT volumes may also differ in the number of B-scans across patients and devices, we further defined the final patient-level metric as normalized SoF by the total number of B-scans in a volume to obtain the final patient-level metric:

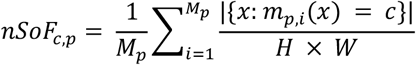

This quantity corresponds to the *mean per B scan fluid area fraction* and was used as the final patient-level measure of fluid burden. We compare model prediction to the ground truth by computing mean absolute error (MAE), which was defined as the absolute difference between the model-predicted and ground-truth reference *nSoF*_*c,p*_ values for each patient and fluid class.

To evaluate agreement between model-derived and annotation-derived fluid burden, we used the intraclass correlation coefficient (ICC), which is widely used to assess agreement of quantitative imaging measurements.^12,13^ We computed ICC using the absolute agreement, two way random effects, single measure specification ICC(2,1).^13^ Our primary metric is the patient-level (aka patient-sum) ICC, which refers to ICC computed on one SoF value per patient, obtained by aggregating fluid burden across all B-scans in that patient’s OCT volume. Due to OCT volumes may differ in the number of B-scans and the number of fluid-containing B-scans across patients, we also computed the ICC at two additional aggregation levels: All-B-scan refers to B-scan-level ICC computed across all B-scans, including those in which the fluid class of interest is absent; Fluid-only-B-scan refers to B-scan-level ICC computed only across B-scans in which the fluid class of interest is present in the reference annotation. For each subject, ICC compares two measurement sources: the ground-truth reference SoF and the model-predicted SoF. ICC values and 95% confidence intervals were computed using *pingouin* (v0.5.11).^14^

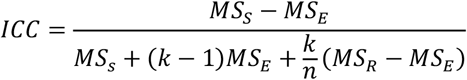

In this formulation, *MS*_*S*_ is the mean square for subjects, *MS*_*R*_ is the mean square for measurement sources, and *MS*_*E*_ is the residual mean square error. Here, *n* denotes the number of subjects and *k* denotes the number of measurement sources. In the patient-level analysis, *n* corresponds to the number of patients. In the B-scan-level analyses, *n* corresponds to the number of B-scans included under the given aggregation level.

#### Image-level qualitative assessment through visualization

We additionally performed qualitative assessment by visually comparing predicted masks with the ground-truth annotations on randomly selected B-scans from both devices (Cirrus and Spectralis). Figure 3 presents these examples with the OCT B-scan, the ground-truth annotation, RFN segmented masks, and VFM segmented masks.

## 3. Results

### 3.1. Pixel-level segmentation performance

Table 1 summarizes pixel-wise segmentation performance (mean ± SD, %) for RetiFluidNet (RFN) and VisionFM (VFM) on RETOUCH across the test folds. Overall, RFN achieved higher performance on most metrics and fluid classes, with the clearest advantages for SRF and PED. Performance on IRF was similar between models for overlap-based metrics: DSC was nearly identical (RFN 80.67 ± 3.14 vs VFM 80.53 ± 1.83), and IoU differed only slightly (RFN 76.30 ± 3.37 vs VFM 76.77 ± 1.92). In contrast, RFN maintained consistently higher BAcc across classes, suggesting more robust class-wise discrimination beyond the dominant background class.

**Table 1.** Pixel-level segmentation performance on RETOUCH (mean ± SD in %, higher is better). ACC: accuracy; BAcc: balanced accuracy; DSC: Dice similarity coefficient; IoU: intersection-over-union. For each metric, the better-performing result is shown in bold.

| Class | ACC | BAcc | DSC | IoU |
| --- | --- | --- | --- | --- |
| **RetiFluidNet (RFN)** |  |  |  |  |
| **Background** | **99.49 ± 0.17** | **91.17 ± 1.44** | **99.74 ± 0.09** | **99.48 ± 0.17** |
| **IRF** | **99.75 ± 0.09** | **94.04 ± 0.29** | **80.67 ± 3.14** | 76.30 ± 3.37 |
| **SRF** | **99.87 ± 0.08** | **96.84 ± 1.32** | **89.83 ± 2.75** | **87.26 ± 3.99** |
| **PED** | **99.85 ± 0.04** | **95.40 ± 1.34** | **86.17 ± 3.49** | **83.95 ± 3.68** |
| **VisionFM (VFM)** |  |  |  |  |
| **Background** | 99.29 ± 0.17 | 87.47 ± 0.07 | 99.63 ± 0.09 | 99.27 ± 0.18 |
| **IRF** | 96.56 ± 2.96 | 88.95 ± 2.22 | 80.53 ± 1.83 | **76.77 ± 1.92** |
| **SRF** | 95.25 ± 3.08 | 90.52 ± 4.84 | 86.95 ± 6.27 | 84.37 ± 7.26 |
| **PED** | 95.47 ± 5.14 | 89.16 ± 5.66 | 83.62 ± 6.18 | 81.52 ± 6.18 |

### 3.2. Patient-level fluid burden quantification performance

**Table 2** reports the comparison between model predictions and the ground-truth reference for patient-level fluid burden estimation using *nSoF* values. Mean predicted *nSoF* and MAE (mean ± SD) relative to the ground-truth reference are shown by device and fluid class.

**Table 2.** Normalized SoF burden (% pixels per B-scan, mean ± SD) for ground-truth reference (GT) and model predictions (Pred), with mean absolute error (MAE), stratified by device and fluid class.

| Device | Class | GT | RetiFluidNet (RFN) |  | VisionFM (VFM) |  |
| --- | --- | --- | --- | --- | --- | --- |
|  |  |  | Pred | MAE | Pred | MAE |
| Cirrus | IRF | $0.40 \pm 0.52$ | $0.43 \pm 0.57$ | $0.03 \pm 0.06$ | $0.55 \pm 0.77$ | $0.17 \pm 0.26$ |
| | SRF | $0.35 \pm 0.77$ | $0.35 \pm 0.77$ | $0.02 \pm 0.03$ | $0.38 \pm 0.84$ | $0.06 \pm 0.10$ |
| | PED | $0.40 \pm 0.90$ | $0.37 \pm 0.78$ | $0.07 \pm 0.16$ | $0.25 \pm 0.56$ | $0.16 \pm 0.39$ |
| Spectralis | IRF | $0.58 \pm 0.73$ | $0.61 \pm 0.78$ | $0.05 \pm 0.06$ | $0.71 \pm 0.97$ | $0.18 \pm 0.27$ |
| | SRF | $0.58 \pm 0.90$ | $0.59 \pm 0.92$ | $0.02 \pm 0.02$ | $0.55 \pm 0.85$ | $0.03 \pm 0.06$ |
| | PED | $0.32 \pm 0.45$ | $0.35 \pm 0.48$ | $0.04 \pm 0.05$ | $0.26 \pm 0.39$ | $0.07 \pm 0.10$ |

Overall, RFN produced predicted *nSoF* values that were close to the ground truth across all fluid classes and devices, with small MAEs and relatively low variability. For Cirrus volumes, RFN showed minimal MAE for IRF (0.03 ± 0.06) and SRF (0.02 ± 0.03), while slightly higher for PED (0.07 ± 0.16). A similar pattern was observed for Spectralis volumes, where RFN MAEs remained small across classes (IRF: 0.05 ± 0.06; SRF: 0.02 ± 0.02; PED: 0.04 ± 0.05). These results indicate that RFN produced fluid burden estimates that were both accurate and stable across devices.

In contrast, VFM showed larger MAE and greater variability for several fluid classes. On Cirrus volumes, VFM substantially larger error for IRF burden (MAE: 0.17 ± 0.26) and PED (0.16 ± 0.39), with noticeably larger standard deviations compared with RFN. On Spectralis volumes, VFM again showed larger error IRF (0.18 ± 0.27) and SRF (0.03 ± 0.06) and PED (0.07 ± 0.10), where SRF error was more comparable between the two models (VFM: 0.03 ± 0.06 vs. RFN: 0.02 ± 0.02). Overall, these results suggest that VFM produced less accurate and less stable patient-level fluid burden estimates than RFN, particularly for IRF and PED.

Figure 2 summarizes agreement, measured by ICC, between model-derived and ground-truth reference estimates of fluid burden quantified by SoF across fluid classes, devices, and the three predefined aggregation levels. RFN (Figure 2A) demonstrated consistently high agreement across fluid classes and devices, with ICC values generally above 0.90 across the three aggregation levels. In contrast, VFM (Figure 2B) showed lower and more variable agreement, most notably on Cirrus for IRF and PED, where ICC values were lower and the 95% confidence intervals were wider than those of RFN. This pattern is consistent with the larger MAE and variability observed for VFM in Table 2, especially for Cirrus IRF and PED, suggesting that VFM yielded less stable fluid burden estimates than RFN for these classes.

**Figure 2.**
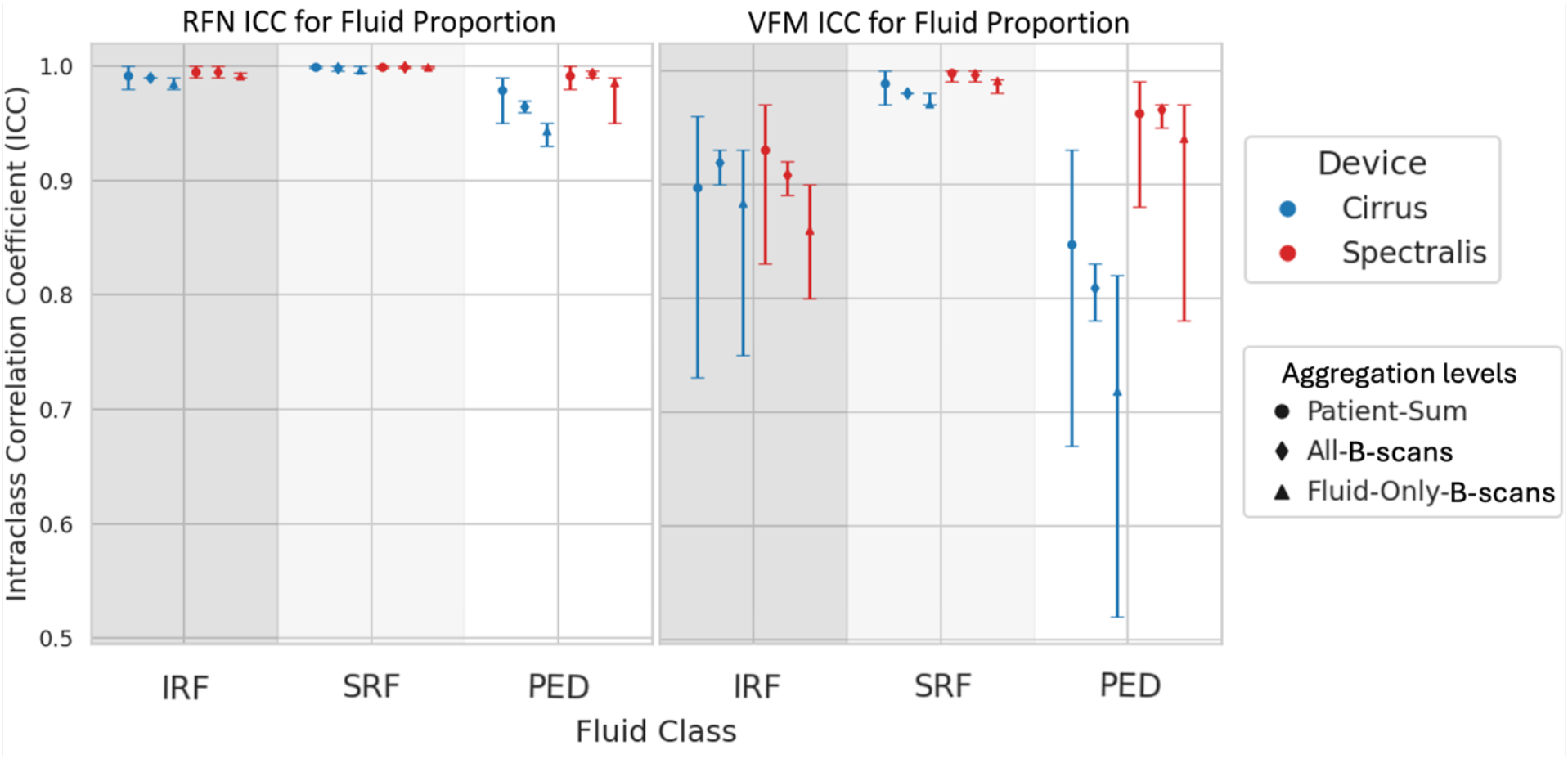
Intraclass correlation coefficients (ICC) between model-derived and ground-truth reference SoF estimates of fluid burden for IRF, SRF, and PED across aggregation levels and devices. **(A)** RetiFluidNet (RFN). **(B)** VisionFM (VFM). Marker shape denotes aggregation level (patient-sum, all-B-scans, fluid-only-B-scans), and color denotes device (Cirrus = blue, Spectralis = red). Error bars correspond to 95% confidence intervals.

### 3.3. Image-level qualitative assessment through visualization

Representative B-scans from Spectralis (top row) and Cirrus (bottom row) are shown in Figure 3. Relative to VFM, RFN more closely aligns with the reference masks for small, focal IRF regions and thin PED layers, with fewer boundary discontinuities. VFM more frequently under-segments low-contrast fluid regions, producing smaller predicted areas than the reference, particularly for subtle IRF and thin PED. Example B-scans were selected to illustrate recurring differences observed across the evaluation set.

**Figure 3.**
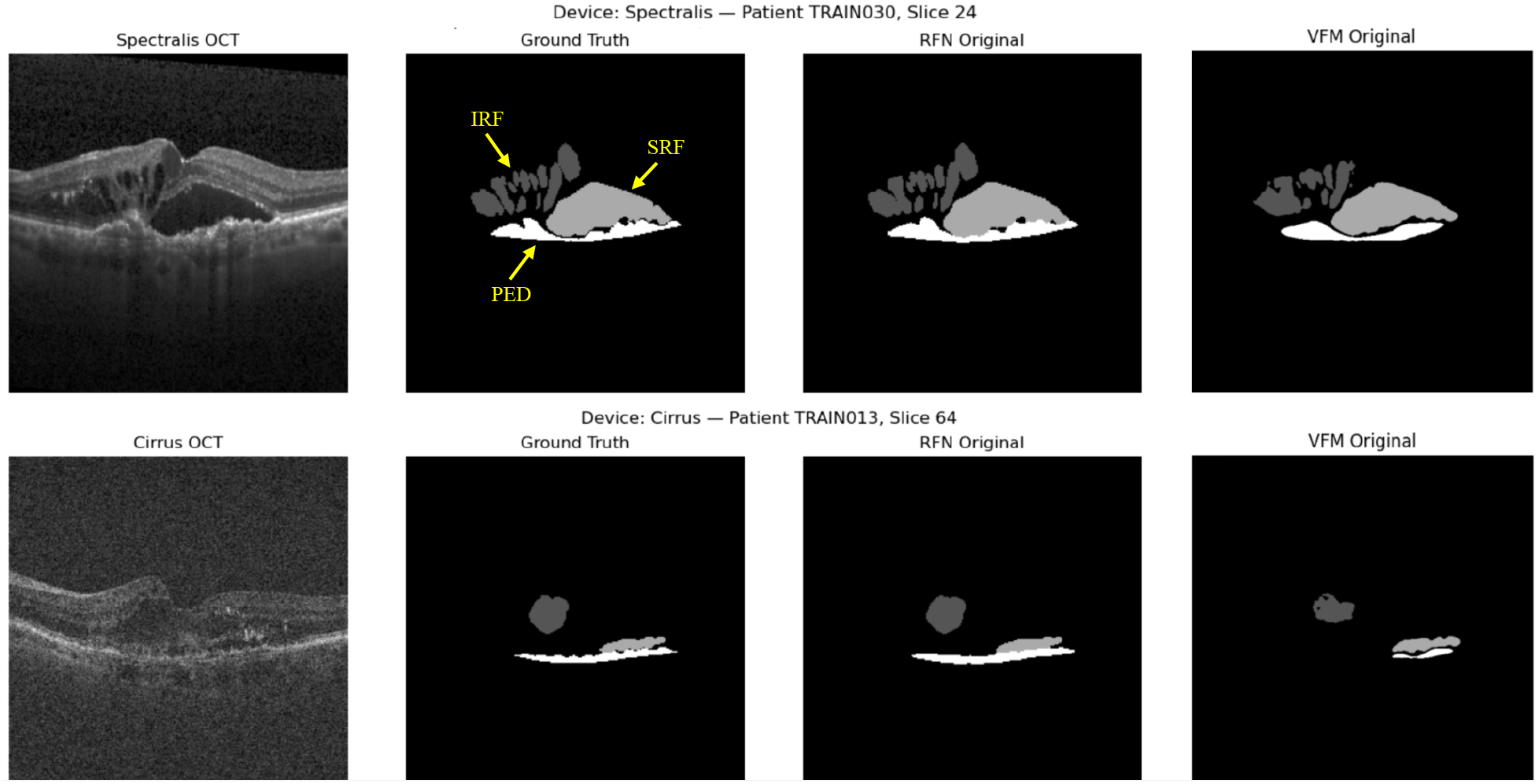
Qualitative comparison of RFN and VFM segmentations on Spectralis (top row) and Cirrus (bottom row) B-scans. From left to right: raw OCT, reference mask, RFN prediction, VFM prediction. In the segmentation masks, dark gray, light gray, and white regions correspond to IRF, SRF, and PED, respectively.

## 4. Discussion

In this head-to-head evaluation on RETOUCH, the task-specific model (RetiFluidNet, RFN) achieved higher pixel-level segmentation performance and more consistent patient-level fluid burden estimates than the foundation model (VisionFM, VFM) for most fluid classes. RFN’s advantage was most evident for SRF and PED, while IRF overlap metrics were comparable across models. Notably, despite similar IRF DSC/IoU, VFM exhibited lower and more variable ICC for IRF SoF-derived fluid burden on Cirrus, indicating that comparable spatial overlap can coexist with less reliable aggregated burden estimates. This DSC/IoU–ICC discrepancy highlights the need to examine systematic bias in burden estimates, device-specific effects, and sensitivity to the chosen aggregation level when models are intended for longitudinal monitoring. To contextualize these findings, we compare them with prior ophthalmic OCT segmentation studies that directly assess foundation-model approaches against task-specific baselines.

Prior work comparing foundation-model approaches with task-specific OCT segmentation models has reported mixed results, with performance depending on the target label set and the extent of OCT-specific adaptation. Consistent with our findings on RETOUCH, SAMedOCT, an OCT-adapted foundation-model approach built on Segmenting Anything (SAM), reported that SAM-based methods can be competitive for OCT biomarker segmentation but generally underperform strong task-specific baselines such as nnU-Net across most settings, and suggested that OCT-specific self-supervised adaptation could help close remaining gaps.^15, 16^ In contrast, studies that explicitly adapt foundation models to OCT report improved segmentation performance on their evaluated datasets. For example, prompt-based volumetric biomarker segmentation methods such as MedSAM 2 (built on SAM 2) can outperform U-Net baselines for biomarkers including IRF and PED, although these interactive settings are not directly comparable to fully automatic segmentation.^17^

More broadly, benchmark work such as MIRAGE emphasizes that domain- and structure-aware pretraining, including retinal structure cues and multimodal retinal data, can improve OCT segmentation quality and robustness, and that foundation-model performance varies substantially across implementations.^18^ VisionFM similarly reports improved OCT segmentation performance relative to U-Net baselines in its benchmark suite, but those results were obtained on different OCT datasets and evaluation settings. Because the benchmark does not specify the same RETOUCH IRF, SRF, and PED labeling setup used here, differences in dataset composition and segmentation targets may partly explain the observed SRF/PED overlap gap and reduced stability in burden estimates.^5^

Overall, these studies place our findings in context by suggesting that task specific models remain strong baselines for fully automatic multi class retinal fluid segmentation, particularly when the downstream goal is reliable quantitative monitoring. Foundation models may require more targeted OCT specific adaptation to achieve comparable reliability in both pixel level segmentation and derived measures of disease burden. From an informatics perspective, these results emphasize that foundation models should be evaluated not only on segmentation overlap metrics but also on the stability of downstream quantitative measures when integrated into clinical monitoring, longitudinal analysis, or decision support pipelines. More broadly, task specific models may remain advantageous when the target task is narrowly defined, annotations are well curated, and precise quantification is required for downstream clinical or research use.

This study has several limitations. First, the evaluation was conducted on a single dataset, which may not capture the full range of clinical variation across institutions, devices, and patient populations. Second, we evaluated one representative model in each category (RFN as task specific and VFM as foundation), so the findings should not be interpreted as generalizable to all task specific or foundation model architectures. Third, VFM was not fully adapted to the SRF and PED segmentation targets used in RETOUCH, including the absence of decoder fine tuning specifically optimized for these classes, which likely contributed to the observed performance gap.

Future work will include class specific adaptation of VFM for SRF and PED, external validation across additional OCT datasets and devices, and evaluation on real world clinical OCT scans. These extensions are important for determining whether automated fluid segmentation can support clinically meaningful longitudinal assessment of disease burden, including detection of worsening or improving fluid patterns during treatment. In practice, more reliable segmentation and burden quantification could support treatment monitoring, response assessment, and cross visit comparison, particularly in diseases such as age-related macular degeneration and diabetic macular edema where retinal fluid dynamics are closely linked to anti-VEGF therapy management.

## 5. Conclusion

A locally trained, task-specific segmentation model (RetiFluidNet) achieved higher retinal fluid segmentation performance than a general-purpose foundation model (VisionFM) and showed more consistent agreement in fluid quantification. This result suggests that task-specific training remains advantageous for OCT fluid segmentation in the evaluated setting and motivates careful validation of foundation models for specialized clinical tasks.

## Data Availability

All data produced can be requested from the organizers of the MICCAI 2017 RETOUCH challenge online at https://retouch.grand-challenge.org/.

## Acknowledgment

This project was partly supported by the National Library of Medicine (NLM) under Award Number R25LM014224.

## References

1. Bommasani R, Hudson DA, Adeli E, et al. On the opportunities and risks of foundation models. arXiv [Preprint]. 2021. arXiv:2108.07258.

2. Zhang S, Metaxas DN. On the challenges and perspectives of foundation models for medical image analysis. Med Image Anal. 2024;91:102996. doi:10.1016/j.media.2023.102996.

3. Lee R, Wong TY, Sabanayagam C. Epidemiology of diabetic retinopathy, diabetic macular edema and related vision loss. Eye Vis (Lond). 2015;2:17. doi:10.1186/s40662-015-0026-2.

4. Rasti R, Biglari A, Rezapourian M, Yang Z, Farsiu S. RetiFluidNet: a self-adaptive and multi-attention deep convolutional network for retinal OCT fluid segmentation. IEEE Trans Med Imaging. 2023;42(5):1413–1423. doi:10.1109/TMI.2022.3228285.

5. Qiu J, Wu J, Wei H, et al. Development and validation of a multimodal multitask vision foundation model for generalist ophthalmic artificial intelligence. NEJM AI. 2024;1(12):AIoa2300221. doi:10.1056/AIoa2300221.

6. Bogunović H, Venhuizen F, Klimscha S, et al. RETOUCH: the retinal OCT fluid detection and segmentation benchmark and challenge. IEEE Trans Med Imaging. 2019;38(8):1858–1874. doi:10.1109/TMI.2019.2901398.

7. Garcea F, Serra A, Lamberti F, Morra L. Data augmentation for medical imaging: a systematic literature review. Comput Biol Med. 2023;152:106391. doi:10.1016/j.compbiomed.2022.106391.

8. Dosovitskiy A, Beyer L, Kolesnikov A, et al. An image is worth 16×16 words: Transformers for image recognition at scale. In: International Conference on Learning Representations (ICLR). 2021. doi:10.48550/arXiv.2010.11929.

9. Zhou J, Wei C, Wang H, Shen W, Xie C, Yuille A, Kong T. iBOT: Image BERT pre-training with online tokenizer. In: International Conference on Learning Representations (ICLR). 2022.

10. Hatamizadeh A, Tang Y, Nath V, et al. UNETR: Transformers for 3D medical image segmentation. In: Proceedings of the IEEE/CVF Winter Conference on Applications of Computer Vision (WACV). 2022. p. 574–584.

11. Cardoso MJ, Li W, Brown R, et al. MONAI: an open-source framework for deep learning in healthcare. arXiv [Preprint]. 2022. arXiv:2211.02701.

12. Xue C, et al. Radiomics feature reliability assessed by intraclass correlation coefficient: a systematic review. Quant Imaging Med Surg. 2021;11(10):4431–4460. doi:10.21037/qims-21-86.

13. Koo TK, Li MY. A guideline of selecting and reporting intraclass correlation coefficients for reliability research. J Chiropr Med. 2016;15(2):155–163. doi:10.1016/j.jcm.2016.02.012.

14. Vallat R. Pingouin: Statistics in Python. J Open Source Softw. 2018;3(31):1026. doi:10.21105/joss.01026.

15. Fazekas B, Morano J, Lachinov D, Aresta G, Bogunović H. SAMedOCT: Adapting Segment Anything Model (SAM) for Retinal OCT. arXiv [Preprint]. 2023. arXiv:2308.09331. doi:10.48550/arXiv.2308.09331.

16. Kirillov A, Mintun E, Ravi N, et al. Segment anything. In: Proceedings of the IEEE/CVF International Conference on Computer Vision (ICCV); 2023. p. 4015–4026.

17. Kulyabin M, Zhdanov A, Pershin A, Sokolov G, Nikiforova A, Ronkin M, et al. Segment Anything in Optical Coherence Tomography: SAM 2 for Volumetric Segmentation of Retinal Biomarkers. Bioengineering (Basel). 2024;11(9):940. doi:10.3390/bioengineering11090940.

18. Morano J, Fazekas B, Sükei E, et al. Multimodal foundation model and benchmark for comprehensive retinal OCT image analysis. npj Digit Med. 2025;8:576. doi:10.1038/s41746-025-01852-3.

19. Pawloff, M., Gerendas, B.S., Deak, G. et al. Performance of retinal fluid monitoring in OCT imaging by automated deep learning versus human expert grading in neovascular AMD. Eye 37, 3793–3800 (2023). 10.1038/s41433-023-02615-8

20. Schlegl T, Waldstein SM, Bogunović H, et al. Fully automated detection and quantification of macular fluid in OCT using deep learning. Ophthalmology. 2018;125(4):549–558. doi:10.1016/j.ophtha.2017.10.031.

